# Trends for the Impact of Cigarette Smoking on Mortality in US States

**DOI:** 10.64898/2026.02.02.26345393

**Authors:** Theodore R. Holford, Jamie Tam, Jihyoun Jeon, Yoonseo Mok, Rafael Meza

## Abstract

**Introduction:** Mortality and smoking rates vary over time across the US. The Cancer Intervention and Surveillance Modeling Network—Lung Working Group (CISNET-LWG) has developed a smoking history generator to describe the effects smoking on health. This work further refines these parameters and quantifies effects on life expectancy

**Methods:** Data from the National Health Interview Survey (NHIS) and the Tobacco Use Supplement to the Current Population Survey (TUS-CPS) were used to estimate smoking history parameters for each state. The age-period-cohort was used in most cases, but an age-cohort mode was used for cessation probabilities. Population mortality data were used to estimate mortality rates for all causes, lung cancer, and non-lung cancer. These were partitioned by smoking status.

**Results:** California and Kentucky are states with more or less aggressive tobacco control. The difference between population cohort life expectancy and life expectancy of never smoker was greater for males than for females, and it was greater in Kentucky than California because of higher smoking rates. These differences decreased with time. Similar result are shown for each state.

**Conclusions:** Variation in smoking parameters and mortality trends vary considerably among states. These show variation in exposure to tobacco smoking and their effects on life expectancy. The Southeast region tends to have greater differences from never smokers because of higher smoking rates. However, there are also other factors affecting mortality rates.

## Introduction

UN Sustainable Development Goals has a primary focus on the reducing premature mortality in all countries.^1^ Within the US, there is considerable variation in mortality among the states, and trends among them varying widely. States in the Far West, Mideast, and New England have steadily increased cohort life expectancy throughout the twentieth century. However, recent cohorts in the Southeast and Southwest have changed little for cohorts born in 1950 to 2000.^2^ The work described here attempts to begin to understand the reasons for these disparities.

It is well documented that cigarette smoking has been a major contributor to health disparities during the twentieth century, having large effects on risk for cancers, especially lung cancer, cardiovascular disease, and respiratory diseases.^3-5^ The Cancer Intervention and Surveillance Modeling Network— Lung Working Group (CISNET—LWG) develops population models for lung cancer and mortality to quantify the effects of cigarette smoking and public health approaches to control mortality. This has included an assessment of the impact of the first Surgeon General’s Report^3^ on the health effects of cigarette smoking.^6^

States vary substantially in smoking rates, with some having more than twice that of others, in 2019.^7^ Utah and California had the lowest smoking prevalence (7.9% and 10.0% respectively), while Kentucky (23.6%) and West Virginia (23.8%) the highest. Smoking prevalence is a result of initiation and cessation trends, factors that can be influenced by a combination of state policies affecting the price of cigarettes and access to programs for successful quitting.^5^ This work was done anticipating that it could be used to evaluate the effect of these smoking trends on health in the U.S., and the work presented here is a first step.

Population data on cigarette smoking has been collected by the National Center for Health Statistics (NHIS) starting in 1965, a nearly 65-year history. These data provide a resource for the entire country, but its design is less successful in capturing the variation among states. The Tobacco Use Supplement to the Current Population Survey (TUS-CPS) provides a larger, state-representative sample that began collecting data in 1992. With a shorter time span of data collection, TUS-CPS is less successful in capturing some aspects of temporal trend. With an analysis that combines these data sources, one can capture the strengths of each survey to quantify both temporal and spatial trends in smoking.

In the final step, the smoking history estimates are used together with the mortality estimates referred to above,^2^ to partition the mortality estimates by smoking status, using an approach developed by Rosenberg, et al.^8^ This yields mortality rates by smoking status. In this paper, we discuss some results for all-cause mortality, but the estimate generated by this work also provides estimates for deaths for lung cancer, and for causes other than lung cancer. In work reported elsewhere, these estimates are used in the smoking history generator (SHG)^6,9,10^ to study alternative questions that may affect lung cancer mortality.

## Methods

To estimate the parameters affecting mortality in a population, it is necessary to quantify the history of smoking history parameters and mortality. SHG will also be used to study the effects of change in the future, which requires estimates that extrapolate the effects that have been seen thus far. This requires quantitative trend estimates of smoking history, mortality, and mortality by smoking status.

### Smoking History

#### Study Sample

NHIS is a nationally representative survey giving self-reported behavior for adult (ages 18+), non-institutionalized US residents from 1965-2024, with 7K-53K in each round (response rate approximately 70%).^11^ The US Census Bureau, as part of the Current Population Survey, administers TUS-CPS, which includes detailed smoking and tobacco data from adults (18+). The survey has been conducted every 3-4 years 1992-2023 in waves that include approximately 250,000 per wave (74% response rate).

Times covered by each survey are shown in Figure 1, giving years surveyed (x-axis) and age (y-axis). The data are cross-sectional, capturing responses at time of survey, but some surveys obtain information on age at initiation and cessation, lengthening the time horizon and providing detail on earlier experiences of the individual. Diagonal lines in Figure 1 show the cohorts that are covered by these data.

**Figure 1.**
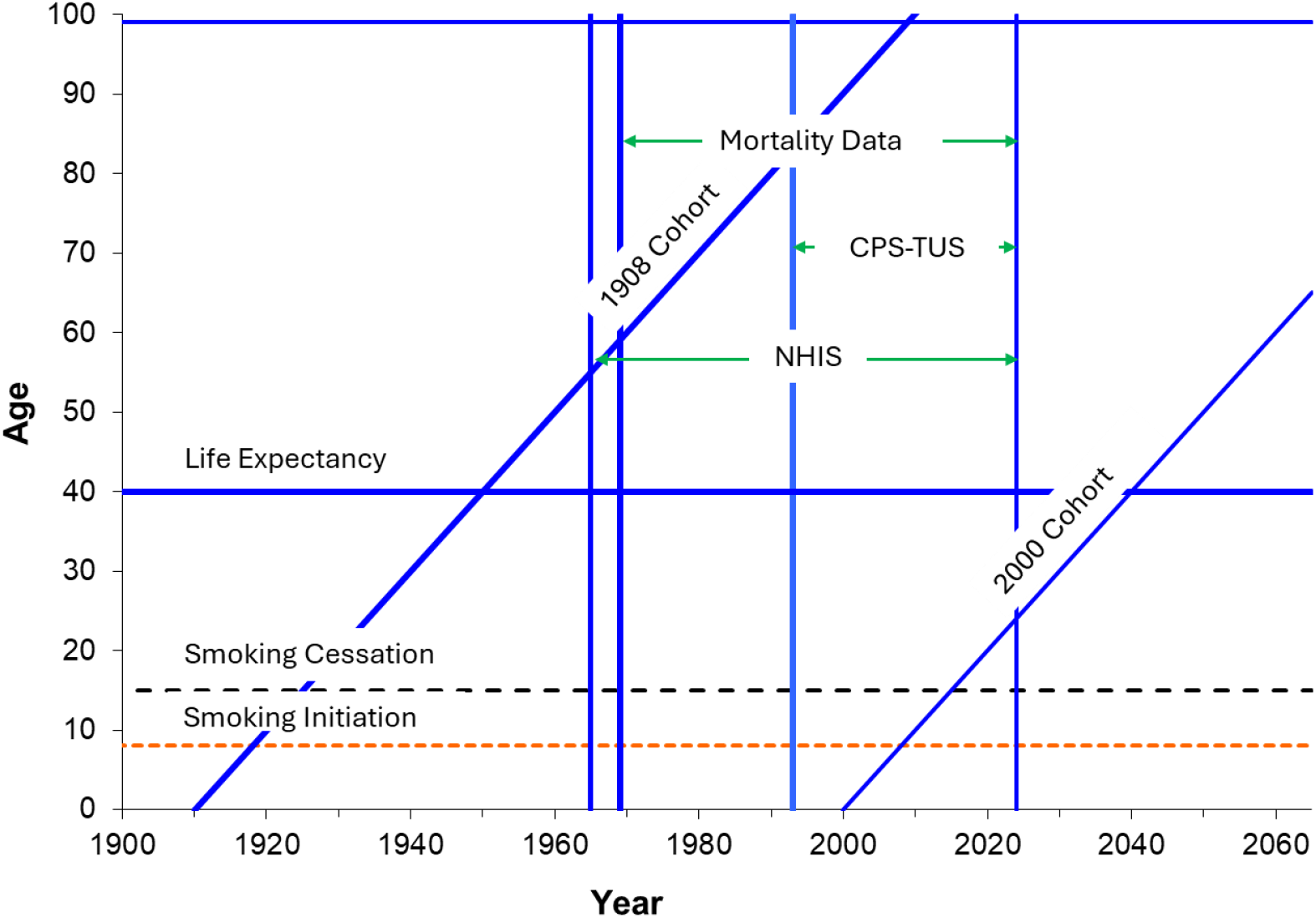
Display of ranges covered by data sources, key starting ages, and cohort limits.

To quantify the experience of a cohort using cross-sectional information results in a gradual decline in prevalence of ever smoking, due to increased mortality in smokers. The later date, i.e., shorter follow-up, for CPS-TUS does not yield a good estimate of this effect for each state. Hence, NHIS is used to estimate this effect for the country, and using this trend provides a correcting calibration of ever-smoker prevalence. The remaining smoking history parameters for each state are derived from TUS-CPC: ever-, current-, and former smoker prevalence, yearly initiation and cessation probabilities, and smoking intensity.

#### Analysis

We first fit an age-period-cohort (APC) model for ages 30+ using NCHS data to obtain an estimate of the age parameter, which estimates the effect of the mortality difference between smokers and non-smokers. This age effect is assumed to be the same for each state, so it is included in the models by using the estimate from NCHS as an offset in an age-cohort model for ever-smoker prevalence in each state for ages 30+. Smoking initiation probabilities were estimated for each state using an APC model, providing estimates for age 8+. Cumulating these estimates provides and estimates of ever-smoking prevalence, but these estimates may not correspond to the direct estimate, in part because of the mortality differences by smoking status that will affect results at the time of survey. Hence, these crude estimated probabilities are calibrated so that the cumulative estimate agrees with the estimate at age 30.

For the cessation probability, an age-cohort (AC) model was used that included interaction parameters. Recent cohort have smoking behaviors that are different from earlier cohorts, and the APC model used in earlier work,^12,13^ does not provide a good fit to the recent data. Current trends exhibit much more rapid increases than before, so extrapolations after the last year of available data approach 1. However, over the range observed, direct estimates are not greater the 0.1, so the estimates are constrained to be no greater than 0.1.

The distribution of smoking intensity categories was estimated using an ordered logistic AC model without interaction terms.

The estimated initiation and cessation probabilities were used to estimate current, former, and never smoker prevalence for each state, using single years for age (8-99), and cohort (1908-2023, and these are extrapolated to 2100).

### Mortality

#### Data sources

Data for estimation of mortality rates by single years of age (0-84) and calendar years were obtained from NCHS, as described earlier.^2^ Figure 1 shows the span of years covered by the mortality data and its relationship the available data on smoking. The online data suppresses frequencies less than 10, resulting in biased estimates if these are simply treated as missing data. To avoid this problem, we obtained permission to use unrestricted data to avoid this difficulty.^14^

#### Analysis

An APC model was fitted to these data, using a Gomperz model for the effect of age 35+, i.e., linear trend for the log rate.^15^ This was accomplished by using constrained cubic splines in which the effect is linear after the final knot.^16^ While the estimates are given for all ages, the focus of this report is the effect of cigarette smoking, which has the greatest effect after age 40.

### Partition Mortality by Smoking History

The final step in estimating parameters needed for the SHG is the estimation of mortality by smoking status. Smoking histories and mortality estimates were used to partition the mortality rates for all-causes and other-causes (not lung cancer), as shown by Rosenberg et al.^8^

## Results

The smoking history parameters have been updated using the more current information described above. These are provided for each state and the country by sex and birth cohorts 1908-2100, and details can be seen for each state on the website for this manuscript (http://xxxx). The trends for prevalence of ever and current smoking show large declines for each state, and the prevalence of never smoking has increased considerably. Reflecting these patterns, smoking initiation probabilities have decreased considerably. For the extension after the 2024 cohort, the cohort parameter is held constant, so that the trends remain the same for the future.

Noticeably different from our earlier analysis are the cessation probability estimates, a result of not using the APC model, but an AC model with interactions. These estimates are considerably higher for more recent cohorts, and the trend is steeper, a pattern that is not well described by the APC model. These estimates have been constrained to be no greater than 0.1 because greater crude estimates for observed data have not been seen thus far. The observed cohorts also show nondecreasing trends, and that is maintained.

The website associated with this work (http://xxxx) gives mortality rate estimates for all causes, lung cancer, and causes other than lung cancer, which are required for population models for lung cancer. It also gives mortality estimates for all causes and for non-lung cancer partitioned by smoking status, and smoking intensity. To understand the overall health impact of cigarette smoking, we focus here on a comparison of all-cause mortality after age 40 for the entire state, and for never smokers. Holford et al^13^ compare smoking parameters for California and Kentucky as states that more and less aggressive in attempting to control cigarette smoking, and the estimates here show the impact of those trends on all-cause mortality. Figure 2 show female cohort life expectancy trends for the population and for never smokers, by state, and country. The difference between the population and never smokers is greater in the earlier cohorts, as would be expected by the decline in smoking. Also, the difference is greater for Kentucky than California, caused by lower smoking rates in the latter.

**Figure 2.**
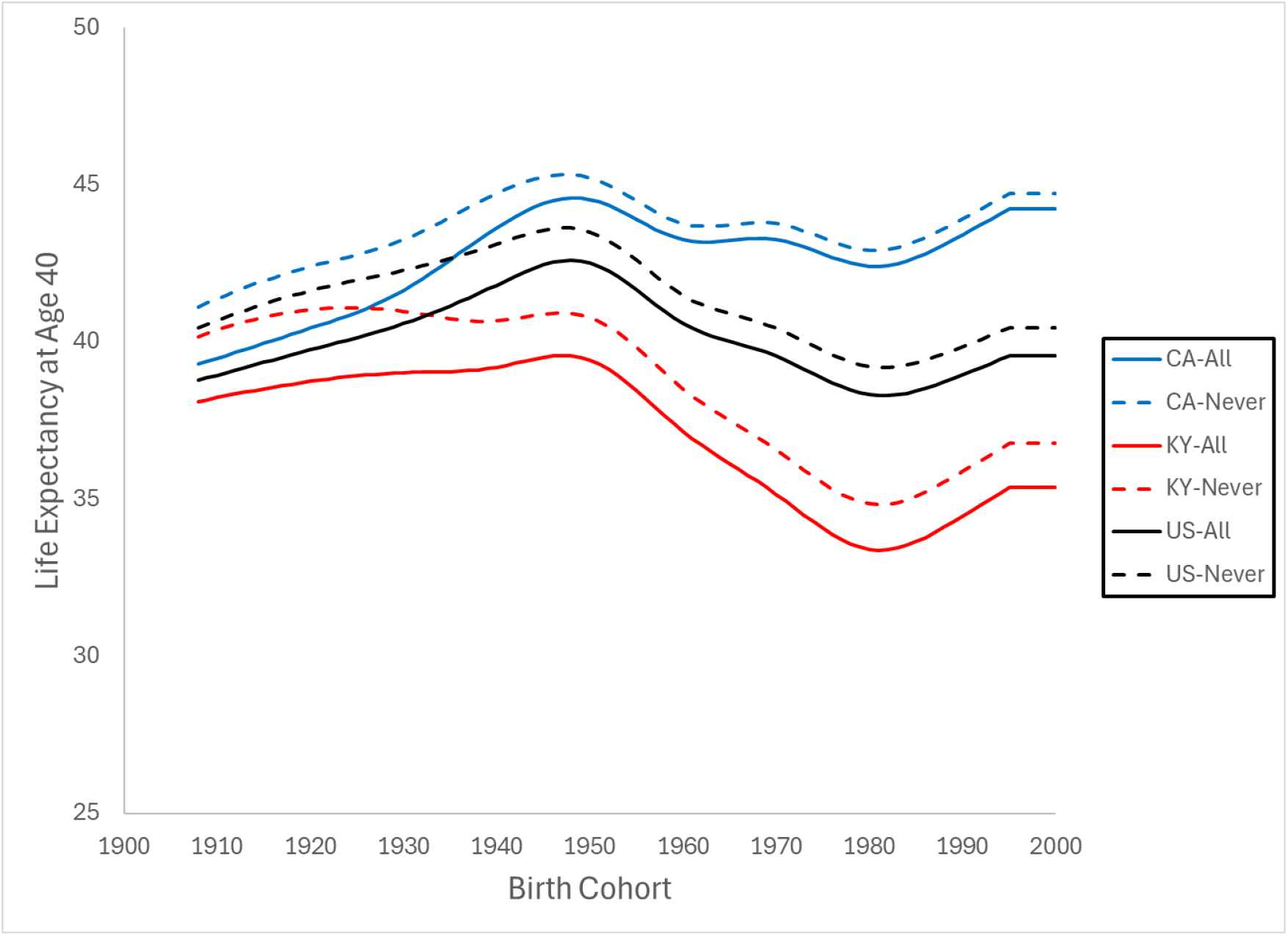
Female life expectancy at age 40 by birth cohort for California, Kentucky and US.

Figure 3 displays trends in cohort life expectancy at age 40 for males. The patterns are broadly similar to those seen in females, with some important distinctions. Males have lower life expectancy than females, and a much larger difference between never smokers and the population. This is especially true in early cohorts, but it remained throughout.

**Figure 3.**
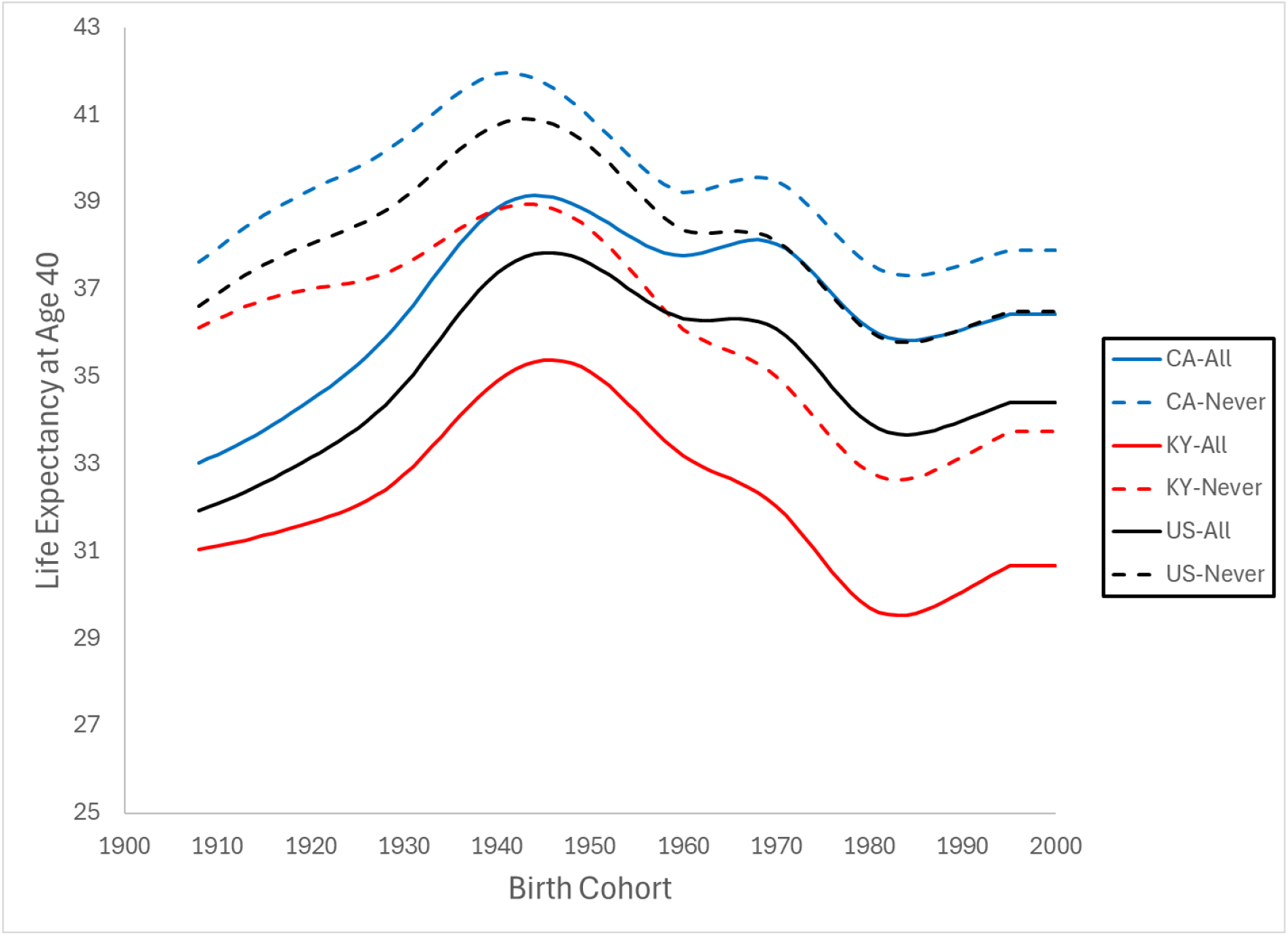
Male life expectancy at age 40 by birth cohort for California, Kentucky and US.

Since 1950, cohort life expectancy has declined for males, particularly in Kentucky (Figure 3). A decline is also seen for females (Figure 2), although smaller. These declines appear to be slightly greater for never-smokers, suggesting that temporal trends in mortality are not a result of changing smoking pattern alone.

Tables 1 and 2 give estimated cohort life expectancies by state for the population, never smokers, and the difference for the 1950 and 2000 cohorts. The difference for Utah is small, about 0.5 years for both sexes, reflecting the predominance of the Mormon population who have low smoking rates. In general, females (Table 1) show a much smaller differences, about one year, than males, which are more varied. The differences for males (Table 2) tend to be highest in the Southeast, about three years or more in 1950 and only slightly less in 2000. The Rocky Mountain and Far West state tend to have the smallest differences between the population estimates and the estimate for never smokers.

**Table 1.** Female life expectancy at age 40 for the population, for never smokers, and the difference for the 1950 and 2000 birth cohorts by state.

| State | 1950 |  |  | 2000 |  |  |
| --- | --- | --- | --- | --- | --- | --- |
|  | All | Never Smokers | Difference | All | Never Smokers | Difference |
| **New England** |  |  |  |  |  |  |
| Connecticut | 44.6 | 45.6 | 1.0 | 40.6 | 41.3 | 0.7 |
| Maine | 43.9 | 45.0 | 1.1 | 45.6 | 46.8 | 1.2 |
| Massachusetts | 44.3 | 45.1 | 0.8 | 40.5 | 41.2 | 0.7 |
| New Hampshire | 43.4 | 44.3 | 0.9 | 37.5 | 38.4 | 0.9 |
| Rhode Island | 44.4 | 45.3 | 1.0 | 44.5 | 45.4 | 0.9 |
| Vermont | 43.6 | 44.5 | 0.9 | 35.7 | 36.6 | 0.9 |
| **Mideast** |  |  |  |  |  |  |
| Delaware | 43.6 | 44.6 | 1.1 | 38.2 | 39.2 | 1.0 |
| District of Columbia | 40.8 | 42.0 | 1.2 | 40.0 | 41.1 | 1.1 |
| Maryland | 43.7 | 44.6 | 0.9 | 40.9 | 41.7 | 0.8 |
| New Jersey | 43.6 | 44.4 | 0.8 | 38.3 | 38.9 | 0.6 |
| New York | 44.4 | 45.3 | 0.9 | 43.3 | 44.0 | 0.7 |
| Pennsylvania | 42.4 | 43.4 | 1.0 | 37.8 | 38.9 | 1.0 |
| **Great Lakes** |  |  |  |  |  |  |
| Illinois | 42.5 | 43.5 | 1.0 | 39.9 | 40.7 | 0.8 |
| Indiana | 40.8 | 42.0 | 1.2 | 36.4 | 37.7 | 1.2 |
| Michigan | 41.6 | 42.8 | 1.2 | 38.1 | 39.3 | 1.1 |
| Ohio | 41.2 | 42.3 | 1.2 | 38.0 | 39.3 | 1.2 |
| Wisconsin | 43.3 | 44.4 | 1.1 | 39.9 | 41.0 | 1.1 |
| **Southeast** |  |  |  |  |  |  |
| Alabama | 39.7 | 40.8 | 1.1 | 35.5 | 36.5 | 1.1 |
| Arkansas | 39.8 | 41.3 | 1.4 | 38.4 | 39.8 | 1.4 |
| Florida | 44.2 | 45.3 | 1.0 | 40.7 | 41.7 | 1.0 |
| Georgia | 41.1 | 42.2 | 1.0 | 38.9 | 39.8 | 0.9 |
| Kentucky | 39.4 | 40.8 | 1.4 | 35.4 | 36.8 | 1.4 |
| Louisiana | 40.3 | 41.6 | 1.3 | 37.5 | 38.8 | 1.4 |
| Mississippi | 39.0 | 40.2 | 1.3 | 35.6 | 36.9 | 1.3 |
| North Carolina | 41.3 | 42.4 | 1.1 | 36.0 | 37.1 | 1.1 |
| South Carolina | 41.1 | 42.1 | 1.0 | 35.5 | 36.6 | 1.2 |
| Tennessee | 39.5 | 40.7 | 1.2 | 33.7 | 35.0 | 1.3 |
| Virginia | 42.5 | 43.4 | 0.9 | 38.9 | 39.7 | 0.8 |
| West Virginia | 39.1 | 40.4 | 1.3 | 31.2 | 32.8 | 1.6 |
| **Southwest** |  |  |  |  |  |  |
| Arizona | 43.2 | 44.0 | 0.8 | 37.7 | 38.4 | 0.8 |
| New Mexico | 42.4 | 43.2 | 0.8 | 32.6 | 33.6 | 1.0 |
| Oklahoma | 39.2 | 40.5 | 1.3 | 36.5 | 37.8 | 1.3 |
| Texas | 41.8 | 42.7 | 0.9 | 42.0 | 42.8 | 0.8 |
| **Rocky Mountain** |  |  |  |  |  |  |
| Colorado | 44.0 | 44.8 | 0.8 | 38.1 | 38.8 | 0.6 |
| Idaho | 42.8 | 43.6 | 0.8 | 37.7 | 38.5 | 0.8 |
| Montana | 43.1 | 44.2 | 1.0 | 36.1 | 37.2 | 1.1 |
| Utah | 43.2 | 43.7 | 0.5 | 39.0 | 39.5 | 0.5 |
| Wyoming | 42.0 | 43.1 | 1.1 | 37.2 | 38.3 | 1.1 |
| **Far West** |  |  |  |  |  |  |
| Alaska | 43.2 | 44.3 | 1.1 | 33.1 | 34.2 | 1.1 |
| California | 44.5 | 45.2 | 0.7 | 44.2 | 44.7 | 0.5 |
| Hawaii | 48.6 | 49.5 | 0.9 | 48.7 | 49.5 | 0.8 |
| Nevada | 42.4 | 43.7 | 1.3 | 41.5 | 42.6 | 1.1 |
| Oregon | 42.8 | 43.6 | 0.9 | 39.2 | 40.0 | 0.8 |
| Washington | 43.4 | 44.4 | 0.9 | 40.3 | 41.2 | 0.9 |
| **United States** | **42.5** | **43.5** | **1.0** | **39.6** | **40.4** | **0.9** |

**Table 2.**
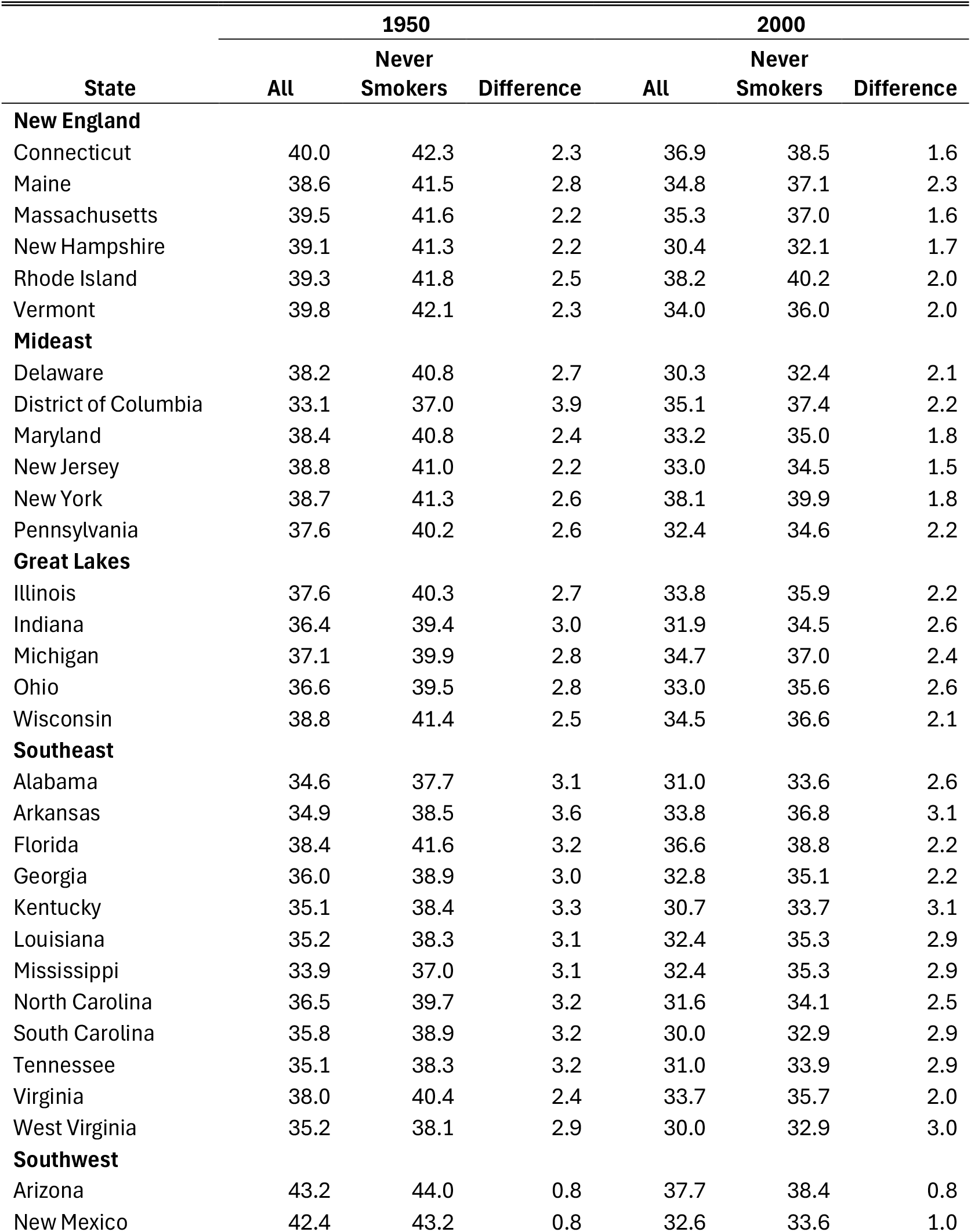
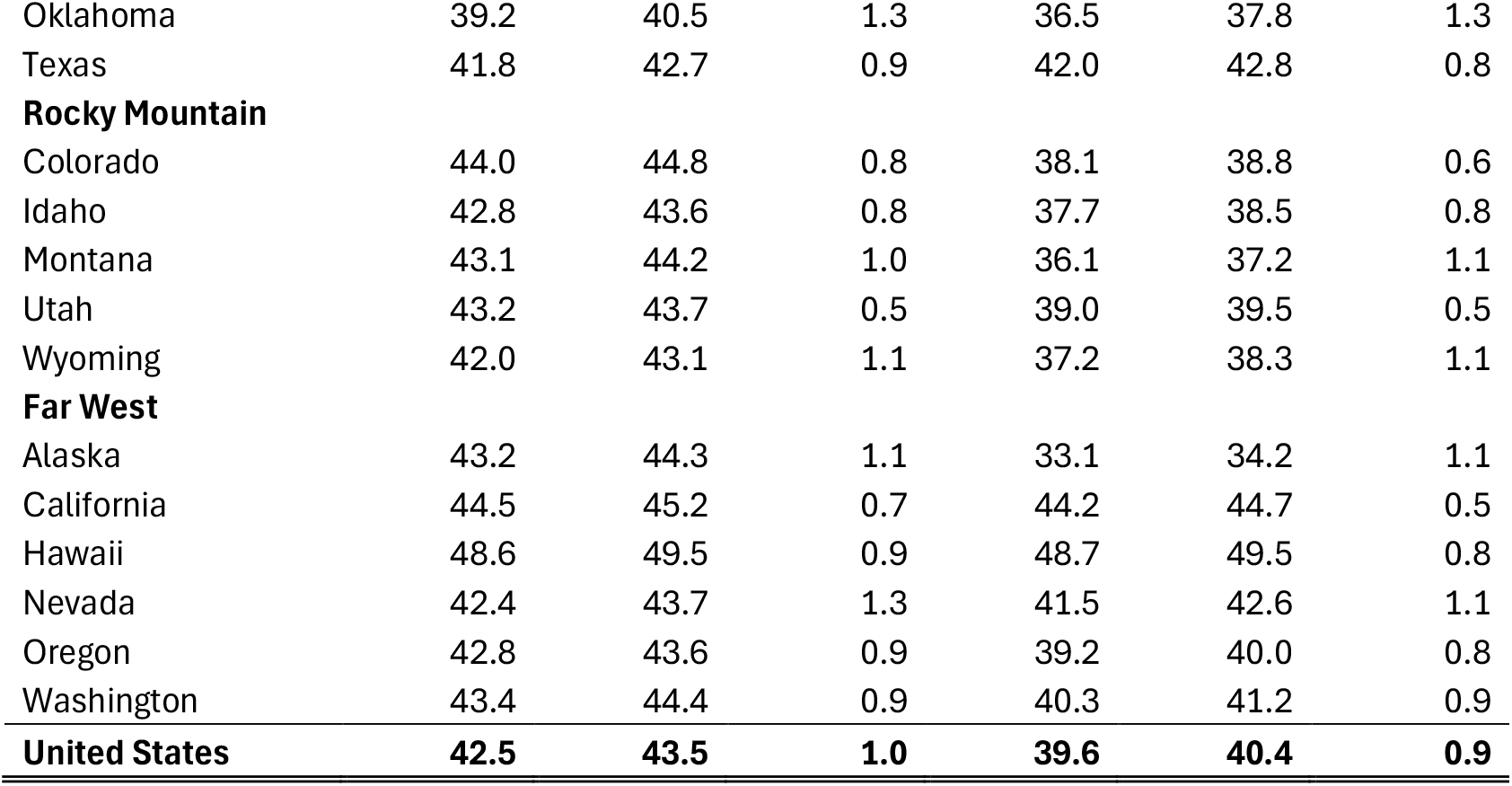
Male life expectancy at age 40 for the population, for never smokers, and the difference for the 1950 and 2000 birth cohorts by state.

The decrease in cohort life expectancy between 1950 and 2000 at age 40 is common among the state. Especially notable is an estimated decline for each sex of about 10 years in Alaska and New Mexico. The District of Columbia estimate increased 2 years for the population, but only 0.4 years for Never smokers.

## Discussion

This work extends earlier work that presents cohort life expectancy trends among US states,^2^ by considering an important factor affecting health, cigarette smoking. We have also showed that smoking trends vary among the states,^13^ and this analysis is an attempt to begin to understand the extent to which this one factor might account for those differences.

The huge decline in cigarette smoking is strongly associated with the change in the primary nicotine delivery method from smoking to vaping. Everyone knew that cigarette smoking was harmful and access to a product that provided the kick without the heavy exposure to carcinogenic tar from burning tobacco was very attractive. Smoking prevalence has plummeted because of much lower initiation probabilities, and much higher cessation probabilities. In the work presented here, cohort effects have been fixed to the value at the last cohort providing data. The trend toward lower initiation and higher cessation shows no sign of letting up so the estimates that we have presented are likely to be higher than what occurs. We have shown that the reduction in exposure to cigarette smoking seen thus far has had very real effects on longevity. These benefits are expected to continue, and they may improve still further. Results shown in the supplement extend the timeframe to future cohorts, which can help to see this progress. However, possible adverse health effects of vaping and other substitutes for cigarette smoking are not well understood at this time so the picture could well change over time.

To quantify the effect of smoking on mortality, we have used the cohort life table to estimate life expectancies for cohorts. Smoking exposure is better understood by looking at the behavior of cohorts because they are essentially based on following a group of individuals over time. The public health policy perspective views a population at a point in time, a cross-section of cohorts at different points in their lives. Cigarette smoking does not begin to have much of an effect on mortality until age 40, so the impact of the effect of smoking behavior on individuals born in 2025 will not begin to be seen until 2065. The effects, of course, continue to grow in time, so it is important that we appreciate that investment in controlling cigarette smoking is mostly an investment for the future.

In these results, we have first considered in some detail the states of California and Kentucky, two states that were also considered in an earlier paper on state-specific smoking trends.^13^ Through the use of taxation and efforts to provide assistance to those trying to stop smoking, California has achieved lower smoking prevalence than Kentucky, a state that has done little or nothing to control tobacco. The harm of cigarette smoking is widely known, and exposure in Kentucky has declined despite the neglect by those in positions of responsibility. The attention of officials can have an additional impact on exposure trends, as can be seen in the comparison of smoking prevalence in these two states.^13^ None of these approaches are completely effective, and it is difficult to remain focused on them. In the work presented here, we have quantified the effect of these differences in smoking trends. The difference between the population life expectancy and that for never smokers is much greater for males than for females, because smoking rates are higher. In addition, the differences are greater for Kentucky than for California, also a result of differences in smoking rates. These differences have consistently declined over time, because smoking rates have declined so that a higher proportion of the population are never smokers or smokers with lower exposure.

In the presentation of all states, Tables 1 and 2, show geographic variability in life expectancy and the effect of smoking. Surprisingly, the changes in life expectancy at age 40 are small or negative for both the overall population and for Never Smokers. This is especially true in the Southeast region of the country. The difference between the population estimates and that of Never Smokers also tends to be greatest in the Southeast, a reflection of high smoking rates in this region. This difference by smoking status has decreased, but it is still substantial in some regions indicating the more work is to be done.

The results presented here are part of an effort to generate parameter estimates that can be used in a smoking history generator developed by the Cancer Intervention and Surveillance Network Lung Working Group (CISNET-LWG). These estimates are projected to the 2100 birth cohort, so that once can assess the impact of changes to current policies affecting lung cancer mortality. These parameters include estimates of trends in smoking prevalence, initiation, cessation, and intensity. Also included are mortality rate estimates for all causes, lung cancer, and causes other than lung cancer. Finally, the estimated parameters include estimated mortality rates for all causes and for causes other than lung cancer by smoking status. These parameter estimates are available for the use of others modeling the effects of smoking on health.

A strength of this work is the use of population data on smoking from NCHS and TUS-CPS, and mortality data by single years of age. This has provided estimates that can be used to quantify the effect of smoking in each state. Health policy is implemented at the state level or even lower. These estimates will be useful to state health departments in their efforts to design the most effective strategies for controlling disease affected by cigarette smoking. The analysis has estimated the effects of age, period, and cohort, using the APC model in some cases. However, the APC model does not working well to describe smoking cessation probabilities in new and old cohorts as part of the same analysis, so this work has also made use of an AC model that includes interaction terms, which provide a better fit to the observed data.

A limitation of these estimates arises from uncertainty in trends going forward. Yearly cessation probabilities have been constrained to be not greater than 0.1, because that is what has been observed thus far. However, it may well not hold in the future.

Another limitation of this analysis arises from the very complex way in which tobacco products are being produced and being used. It is challenging to develop useful models for these behaviors and to quantify their effects on health. In addition, the modelling framework used here overly simplifies the transitions from never to current to former smokers. The final transition can be very difficult, and it often takes multiple attempts.

## Data Availability

All data in this study were obtained from the National Center for Health Statistics and the Tobacco Use Survey-Current Population Survey. The mortality data required permission from the Nation Center for Health Statistics, which was obtained.

https://www.cdc.gov/statesystem/cigaretteuseadult.html

https://www.cdc.gov/nchs/nvss/deaths.htm

## References

1. Norheim OF, Jha P, Admasu K, et al. Avoiding 40% of the premature deaths in each country, 2010-30: review of national mortality trends to help quantify the UN Sustainable Development Gpa; for health. Lancet. 2015;385:239–252. doi:10.1016/S0140-6736(14)61591-9

2. Holford TR, McKay L, Tam J, Jeon J, Meza R. All-cause mortality and life expectancy by birth cohort across US States. JAMA Network Open. 2025;8(4):e257695. doi:10.1001/jamanetworkopen.2025.7695

3. Surgeon General’s Advisory Committee on Smoking and Health. Smoking and Health: Report of the Advisory Committee to the Surgeon General of the Public Health Service. U.S. Department of Health, Education, and Welfare, Public Health Service, Office of the Surgeon General; 1964:

4. U.S. Department of Health and Human Services. The Health Consequences of Smoking-50 Years of Progress. U.S. Departmentof Health and Human Services, Centers for Disease Control and Prevention,National Center for Chronic Dease Prevention and Health Promotion, Office on Smoking and Health; 2014.

5. U.S. Department of Health and Human Services. Smoking Cessation. A Report of the Surgeon General. U.S. Department of Health and Human Services, Centers for Disease Control and Prevention, National Center for Chronic Disease Prevention and Health Promotion, Office on Smoking and Health; 2020.

6. Holford TR, Meza R, Warner KE, et al. Tobacco control and the reduction in smoking-related premature deaths in the United States, 1964-2012. J Am Med Assoc. 2014;311(2):164–171. doi:doi: 10.1001/jama.2013.285112

7. Centers for Disease Control and Prevention. Map of Current Cigarette Use among Adults. U.S. Department of Health & Human Services. Accessed January 5, 2021, 2021. Https://www.cdc.gov/statesystem/cigaretteuseadult.html

8. Rosenberg MA, Feuer EJ, Yu B, et al. Cohort life tables by smoking status, removing lung cancer as a cause of death. Risk Anal. 2012;32:S25–S38.

9. Holford TR, Levy DT, McKay LA, et al. Patterns of Birth Cohort–Specific Smoking Histories, 1965–2009. Am J Prev Med. 2014;46(2):e31–e37. doi:10.1016/j.amepre.2013.10.022 External Link

10. Jeon J, Holford TR, Levy DT, et al. Smoking and Lung Cancer Mortality in the United States from 2015 to 2065. Ann Intern Med. 2018;169(10):684–693. doi:10.7326/M18-1250

11. Carlson SA, Densmore D, Fulton JE, Yore MM, Kohl HW. Differences in physical activity prevalence and trends from 3 U.S. surveillance systems: NHIS, NHANES, and BRFSS. Journal of Physical Activity and Health. 2009;6(1):S18–S27.

12. Holford TR, Levy DT, McKay LA, et al. Patterns of Birth Cohort–Specific Smoking Histories, 1965–2009. Am J Prev Med. 2014;46(2):e31–e37. doi:doi: 10.1016/j.amepre.2013.10.022

13. Holford TR, McKay L, Jeon J, et al. Smoking Histories by State in the U.S. Am J Prev Med. 2023;64(4S1):S22–S31.

14. National Center for Health Statistics. Data from: Detailed Mortality - Limited Geography, 1999–2020. 2022.

15. Gompertz B. On the nature of the function expressive of the law of human mortality, and on a new mode of determining the value of life contingencies. Philosophical Transactions of the Royal Society of London. 1825;115:513–583.

16. Durrleman S, Simon R. Flexible regression models with cubic splines. Stat Med. 1989;8:551–561. doi:doi: 10.1002/sim.4780080504

